# Intraindividual Variability Differentiated Older Adults with Physical Frailty and the Role of Education in the Maintenance of Cognitive Intraindividual Variability

**DOI:** 10.1101/2024.05.16.24307475

**Authors:** Jingyi Wu, Jinyu Chen, Juncen Wu, Chun Liang Hsu

## Abstract

**Objectives:** Physical frailty is associated with increased risk of cognitive impairment. However, its impact on sustained cognitive processing as evaluated by intraindividual variability (IIV), and factors beneficial to IIV in physically frail older adults remain unexplored. This study aimed to quantify differences in IIV between older adults with and without physical frailty, and examine whether education facilitated maintenance of IIV.

**Methods:** This cross-sectional study included 121 community-dwelling older adults 65-90 years with/without physical frailty (PF and non-PF; n=41 and n=80 respectively). Physical frailty was determined via Short Physical Performance Battery. Dispersion across the seven components of the Montreal Cognitive Assessment (MoCA) was computed to ascertain IIV. Multivariate analysis of covariance was used to determine group differences in total score and IIV. Four moderation models were constructed to test the effects of education on age-total score and age-IIV relationships in PF and non-PF.

**Results:** Compared with non-PF, PF showed greater IIV (*p* = .022; partial η² = 0.044). Among PF, education moderated age-total score (R-sq = 0.084, F = 5.840, *p* < 0.021) and age-IIV (R-sq = 0.101, F = 7.454, *p* = 0.010) relationships. IIV increased with age for those with five years (β = 0.313, *p* = 0.006) or no formal education (β = 0.610, *p* = 0.001). Greater than seven years of education (β = 0.217, *p* = 0.050) may be required to maintain IIV at older age.

**Conclusion:** IIV may be a sensitive method to differentiate physically frail older adults. Additionally, perceived cognitive benefits of education may be dependent on physical functioning.

## Introduction

Physical frailty is a clinical syndrome characterized by diminished strength, endurance, and reduced physiological function in individuals (1). It is a prevalent geriatric condition that impacts 11% of older adults over 65 years old worldwide (2, 3). Physically frail older adults are more vulnerable to external and internal stressors than non-frail older adults, leading to a significantly increased risk for cognitive impairment, a loss of independent living, hospitalization, and death (4).

Physical frailty is significantly associated with mild cognitive impairment (MCI) and dementia (5–7). Specifically, a longitudinal study involving 2,305 older adults over the age of 70 years found that those with physical frailty showed significantly greater cognitive decline over 5 years, compared with older adults without physical frailty (8). Likewise, results from the Rush Memory and Aging Project showed that each increase of one point in frailty score (computed as a composite score based on grip strength, timed walk, body composition, and fatigue) at baseline was correlated with 60% increase in the risk of subsequently developing MCI after adjusting age, sex, and education (9).

While MCI is commonly identified via evaluating the total score on the Montreal Cognitive Assessment (MoCA) (10) or the Mini-Mental State Examination (11), such static representation of cognitive function only reflect one aspect of an individual’s cognitive capabilities; wherein the ability to sustain cognitive processing across different cognitive domains (12) is another important facet of cognition.

Intraindividual variability (IIV) is a construct of sustained cognitive processing (13). Distinct from cognitive performance identified by total scores, IIV is measured by calculating the variability or fluctuations across different cognitive domains within a standardized neuropsychological test, or across different tests (14). The three most well-established indices of IIV include: (1) variability within a person on a single test over long periods of observation (e.g., across multiple years), referred to as *intraindividual changes*; (2) variability within a person on repeated trials of a single test on one occasion or over multiple occasions (i.e., hours, days, or weeks), referred to as *inconsistency*; and (3) variability within a person on a single occasion across multiple cognitive domains, referred to as *dispersion* (15, 16).

Notably, IIV has been shown to be linked with neurogenerative disorders (17). For instance, study showed significant differences in IIV as indexed by inconsistency between older adults who were cognitively intact vs. those who were cognitively impaired or had dementia (18). Burton et al. (19) found that individuals with Alzheimer’s Disease exhibited greater IIV as indexed by inconsistency compared to those with Parkinson’s disease. Moreover, studies demonstrated that greater IIV, as indexed by dispersion, was associated with increased risk of MCI and dementia (20, 21). While the majority of the current literature assessed IIV through inconsistency, a recent meta-analysis concluded that compared with inconsistency, dispersion is a more sensitive index of IIV in detecting psychiatric and neurological conditions. Nevertheless, evidence on whether physically frail older adults demonstrate poorer sustained cognitive processing remains unexplored (17).

Years of education have been widely regarded as a protective factor that mitigated cognitive decline (22). For example, compared with older adults with more than two years of education, those with no formal education were ten times more likely to develop cognitive impairment (23). A systematic review concluded that lower education level was associated with an increased risk of AD and dementia (24). Importantly, Alley et al. (25) suggested that an average of 16 years of education can slow the rate of decline in global cognitive function in older adults relative to their counterparts with four years of education. However, no studies to date have identified the number of years of education required to prevent decline in sustained cognitive processing in physically frail older adults who are at significantly greater risk for cognitive impairment and dementia.

Therefore, this study aimed to address two primary questions: (1) compared with older adults without physical frailty, whether older adults with physical frailty exhibit greater IIV; and (2) whether education can moderate the negative effects of aging on IIV in older adults with physical frailty. We hypothesized that older adults with physical frailty would display greater IIV, and years of education would significantly moderate the association between age and IIV as indexed by dispersion.

## Materials and Method

### Study Design and Participants

This was a cross-sectional study involving a total of 121 physically frail (PF) (n=41) and non-PF community-dwelling older adults (n=80) between the age of 65-90 years. Participants were recruited from local community centers and non-government organizations (i.e., institutional research hubs) using posters and advertisements through email, and mobile phone applications (i.e., WhatsApp) from August 2023 to February 2024. Data were collected at the Hong Kong Polytechnic University from September 2023 to March 2024. Ethical approval was obtained from the Institutional Review Board (IRB) of The Hong Kong Polytechnic University (HSEARS20230131001). Written informed consent was obtained for all study participants enrolled into the study.

### Descriptors

Age of participants was recorded in years. Height and weight were measured in units of centimeters (cm) and kilograms (kg).

### Physical Frailty Characterization

As recommended by the European Medicines Agency (EMA) (26–28), physical frailty was evaluated by the Short Physical Performance Battery (SPPB) (29). The SPPB is a validated standardized test for evaluating general mobility and balance (30). The SPPB has been shown to have excellent psychometric properties when used in the elderly to discriminate between frail and non-frail older adults (31). The test consists of three subscales (balance test, 4-meter walk at usual pace, and timed chair sit-to-stand test). During the balance test, participants were first instructed to stand with their feet together, then moving into semi-tandem and full-tandem positions. Participants were asked to maintain each position for 10 seconds. During the 4-meter walk, participants were asked to walk four meters at comfortable speed with an initial and terminal spatial buffer of 1-meter to remove potential effects from acceleration/deceleration. The 4-meter walk was performed twice, and the averaged time taken to perform the test was calculated. During the chair sit-to-stand test, participants were instructed to fold their arms across their chest, stand up from a sitting position on the designated chair, and return to the seated position as quickly as possible five times. The time was recorded from the initial sitting position to the final stand position. Each subscale is scored with a maximum of four points for a total of 12 points, with a higher score indicating better general mobility. A score of ≤ 9/12 is indicative of physical frailty (32).

### Inclusion criteria

Older adults were included if they: (1) were between 65 and 90 years old; (2) were living in the community; (3) were able to ambulate up to four meters with or without assistive devices; (4) were able to provide written informed consent; (5) had access to the internet.

### Exclusion criteria

All participants who met any of the following criteria were excluded: (1) diagnosed with central nerve system diseases that substantially affect cognitive function (i.e., dementia, Parkinson’s, Alzheimer’s disease, Amyotrophic lateral sclerosis, and stroke); (2) living in nursing home or other care facilities/institutions; (3) taking psychotropic medication that influences cognitive and physical function; (4) unable to understand, speak, and read Cantonese/Chinese/English.

### Primary Outcome Measures

#### Education

Education level attained by each study participant was recorded in units of years.

#### Cognitive Function

The Hong Kong version of the MoCA was administered (33). The MoCA is a validated measure of global cognitive function with high specificity and sensitivity in identifying individuals with MCI (10). The MoCA is comprised of seven domain-specific components (visual-spatial, naming, attention, language, abstraction, delay, and orientation). An additional point was given to participants who received ≤ 12 years of education (10). The total score ranges from 0-30 points, with a score ≥ 26/30 indicating unimpaired global cognition (10), 18-25/30 indicating MCI, and < 18/30 indicating signs of dementia (34). The MoCA was used to compute IIV.

#### Computation of Intra-Individual Variability

Computation of IIV as indexed by dispersion was performed through four steps. For clarity, within the context of the present paper we refer to IIV as IIV indexed by dispersion. First, each raw subset score of MoCA was Z-transformed separately according to the distribution of entire older adults (n = 121) (Eqs. (1)) (35). Second, the sum of each participant’s z-transformed score for each of the seven components of MoCA - *A*_i_was calculated by Eqs. (2) (20). Third, the variability in each of the seven components of MoCA was calculated by Eqs. (3) (20). Finally, the square root of the sum of variability in the seven components of MoCA was calculated by Eqs (4) to derive the amount of dispersion across the seven components of MoCA (36).

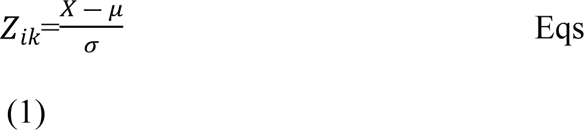

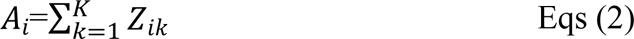

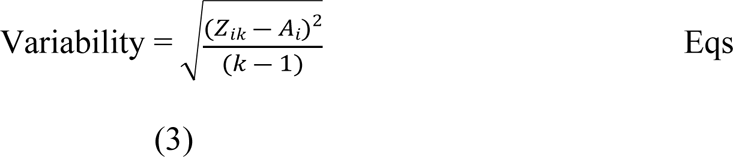

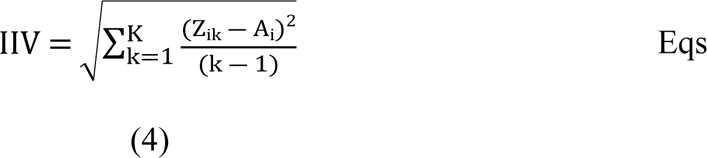

*Z*_ik_was the *k*th cognitive test score for the ith individual. μ was the mean value of all tests. X was the raw score of each test. σ represented the standard deviation of all tests. K represented the number of cognitive tests. *A*_i_ was the individual’s sum Z transformed score based on the number of tests. Note that the number of tests reflects the number of components within the MoCA.

### Statistical Analysis

R software v.4.3.2 (R Development Core Team, 2010) was used to perform all statistical analyses. First, the Shapiro-Wilk’s test was used to ensure the normality of the distribution of all variables (37). Independent t-tests, Mann-Whitney U tests, and chi-squared tests (for ratio and nominal data, respectively) were performed to compare the differences in demographic variables and clinical parameters between the two groups. Mahalanobis distance was used to detect multivariable outliers.

Multivariate Analysis of Covariance (MANCOVA) was used to determine whether there were differences between groups in MoCA total score and IIV adjusting for the effects of age (38). The statistics significance level was set at *p* < 0.05, with correction for multiple comparisons via Bonferroni adjustments at *p* < 0.025. The effect size of the difference between groups was calculated by partial eta squared, where 0.01, 0.06, and 0.14 represented small, medium, and large effect sizes, respectively (39).

Moderation analyses were conducted using the PROCESS macro in R software version 4.3.2 (40). Four separate moderation models were constructed (Fig 1). Two separate models were constructed (one for the non-PF group and one for the PF group) to test the direct effect of age on the MoCA total score and investigate the moderation effect of education on the association between age and the MoCA total score. Two additional models were constructed (one for the non-PF group and one for the PF group) to test the direct effect of age on IIV and investigate the moderation effect of education on the association between age and IIV. The bias-corrected bootstrap confidence intervals were calculated to test the significance of the interaction effect and to control for the possibility of the non-normal distribution of sampling (41). The bootstrap estimates were based on 10,000 bootstrap samples. The interaction effects were considered significant if the upper and lower limits of the 95th percentile CI did not contain zero. To further understand the nature of this interaction, the conditional effect of age (simple slopes) on MoCA total score and IIV was estimated, independently, at five levels of the values of the moderator (i.e., years of education): very low (i.e., 10^th^ percentile), low (i.e., 25^th^ percentile), middle (i.e., 50^th^ percentile), high (i.e., 75^th^ percentile), and very high (i.e., 90^th^ percentile). We also utilized the Johnson-Neyman technique to identify the values of the moderator (i.e., years of education) where the slope of the predictors (i.e., age) is statistically significant (40). The statistical significance level was set at *p* < 0.05 for all tests.

**Fig 1.**
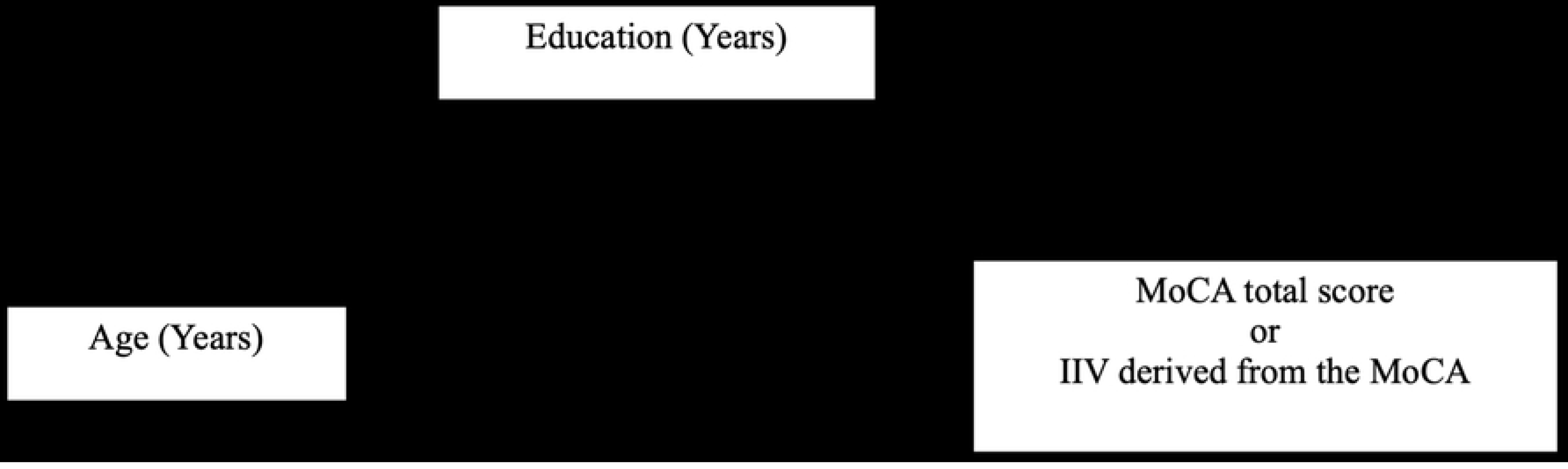
Moderation Model.

## Results

### Participants

Study participants were stratified into the PF (n=41) or the non-PF (n=80) groups. After removing one outlier (i.e., in MoCA and IIV) from the PF group, the PF group included 40 older adults. Participant characteristics are reported in Table 1. The PF group was older and shorter in height than the non-PF group (*p<*0.01 and *p<*0.05 respectively). No other significant differences in characteristics were observed (*p>*0.05). There were trend-level differences in the number of males and females between the two groups (*p<*0.08) (**Table 1**).

**Table 1.** Participant Characteristics.

|  | Physically Frail Group | Non-Physically Frail Group |  |
| --- | --- | --- | --- |
| Demographic data | (n=40) | (n=80) | *p* value |
|  | Mean (SD) | Mean (SD) |  |
| Age (years) | 76.250 (4.960) | 72.938 (3.931) | < 0.001* |
| Sex (M/F) | 9/31 | 33/47 | 0.068 |
| Education (years) | 8.225 (5.512) | 11.900 (4.040) | < 0.001* |
| Weight (kg) | 56.799 (10.389) | 59.125 (9.926) | 0.320 |
| Height (cm) | 156.087 (8.282) | 160.666 (8.222) | 0.013* |
| SPPB (max 12 points) | 7.325 (1.439) | 11.188 (0.731) | < 0.001* |
| MoCA total score (max 30 points)<sup>a</sup> | 22.642 (0.493)<sup>a</sup> | 26.192 (0.342)<sup>a</sup> | < 0.001*<sup>a</sup> |
| IIV<sup>a</sup> | 3.945 (0.419)<sup>a</sup> | 2.725 (0.290)<sup>a</sup> | 0.022*<sup>a</sup> |

\**p* < 0.05
M: male; F: female; SPPB: Short Physical Performance Battery; MoCA: Montreal
Cognitive Assessment; IIV: intraindividual variability as indexed by dispersion.
a. Covariate was evaluated at age = 74.084; mean (SE); SE: standard error.

### MoCA Total Score and IIV between the two groups

The mean MoCA total score, mean IIV, and the group differences adjusted for age are reported in **Table 1**. We found that compared with the non-PF group, the PF group had a significantly lower MoCA total score (mean difference = −3.550; *p* < .001; partial η² = 0.221). Further, compared with the non-PF group, the PF group showed significantly greater IIV (mean difference = 1.220; *p* = .022; partial η² = 0.044). Fig 2 illustrates the differences in the MoCA total score and IIV between non-PF and PF groups.

**Fig 2.**
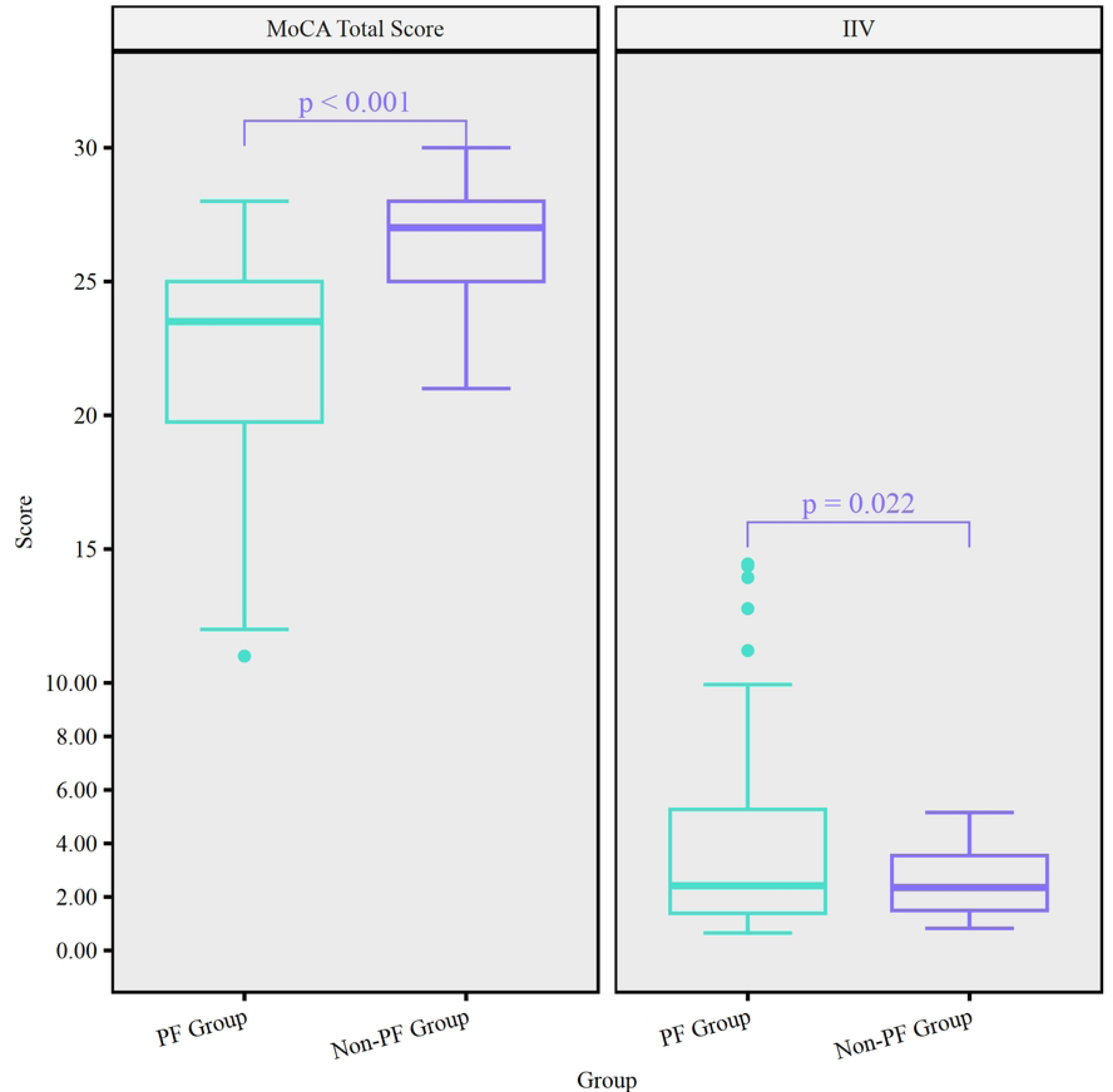
Differences in MoCA total score and IIV between PF group and non-PF group.

### Moderation Effects of Education on the Association between Age and MoCA Total Score

Years of education had a significant moderation effect on the association between age and MoCA total score in the PF group. The overall model was statistically significant (R-sq = 0.482, F = 11.146, *p* < 0.001) (Table 2). Specifically, age exerted a significant negative main effect on the MoCA total score (β = −0.671, SE = 0.182, *p* = 0.001, 95% CI [−1.041, −0.301]). We found a significant two-way interaction between education and age in the PF group (β = 0.058, SE = 0.024, t = 2.417, *p* = 0.021, 95% CI [0.009, 0.107]; Fig 3A) (Table 2), accounting for 8.410% of the variance in the MoCA total score (F = 5.840, *p* = 0.021).

**Fig 3.**
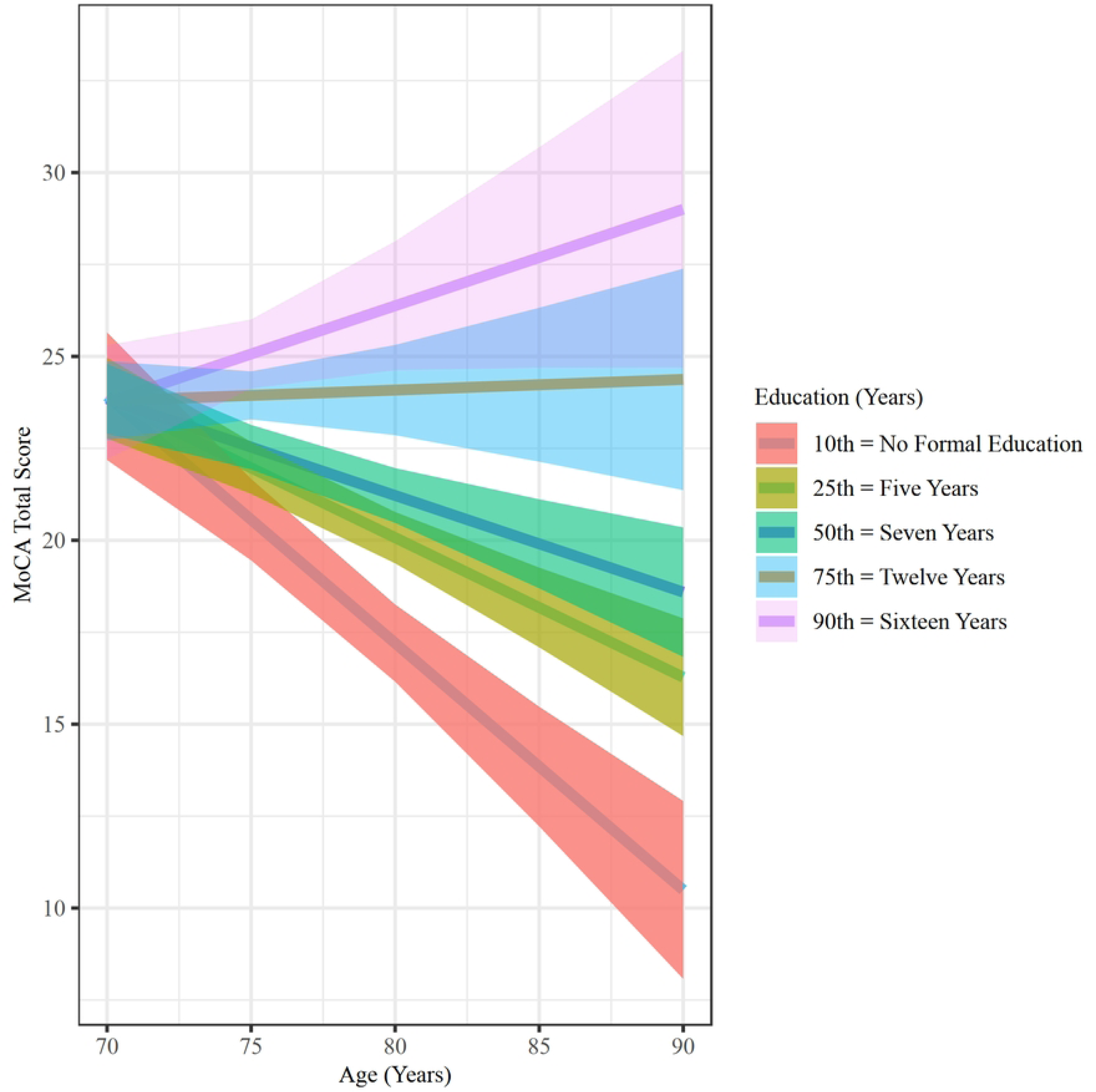
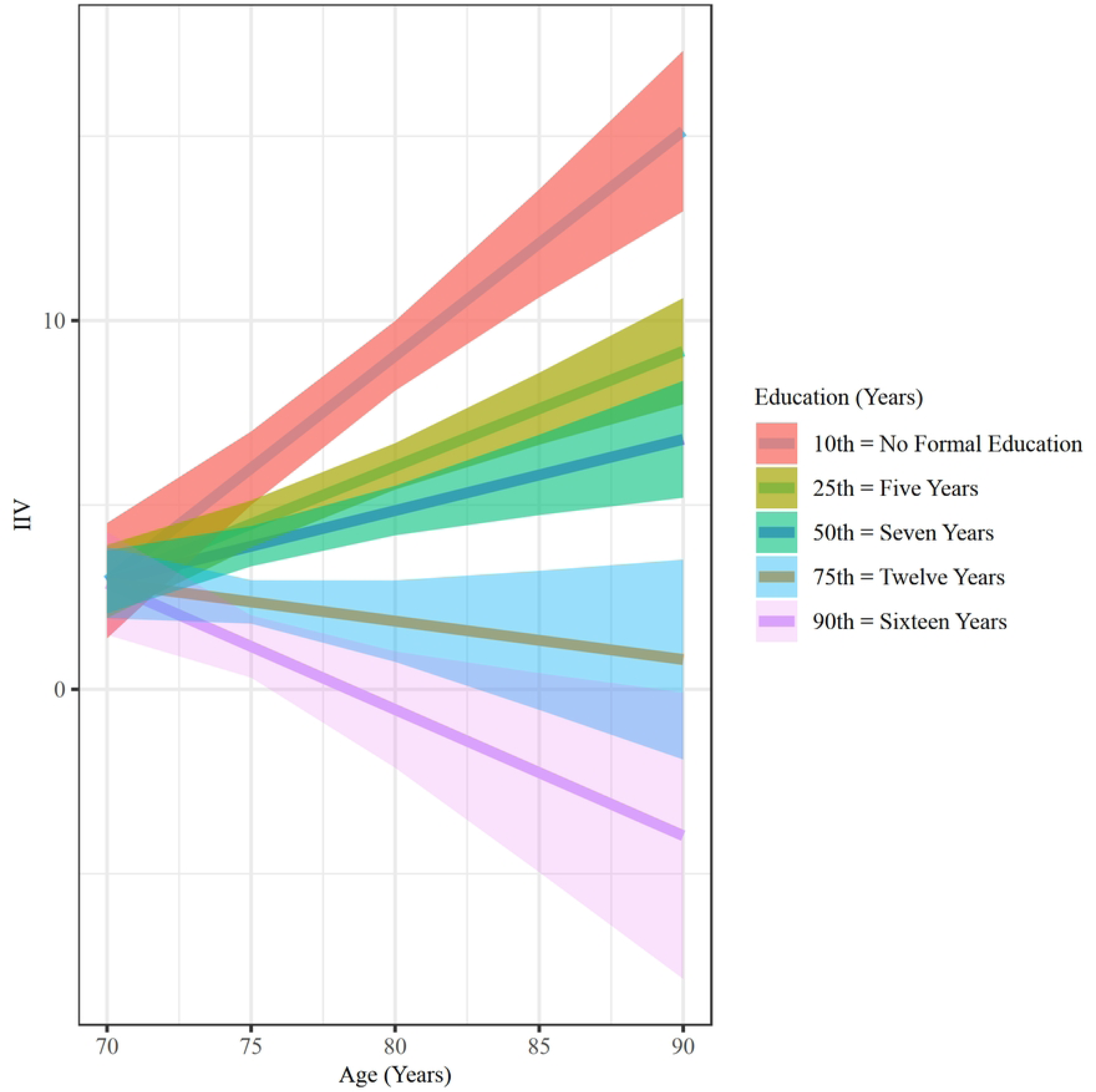
Moderation Effects of Education on the Association between age-MoCA in the Physically Frail Group. (A) the association between age and MoCA total score. (B) Jonnson-Neyman plot for visualizing the moderating effect of education between age and MoCA total score.

**Table 2.**
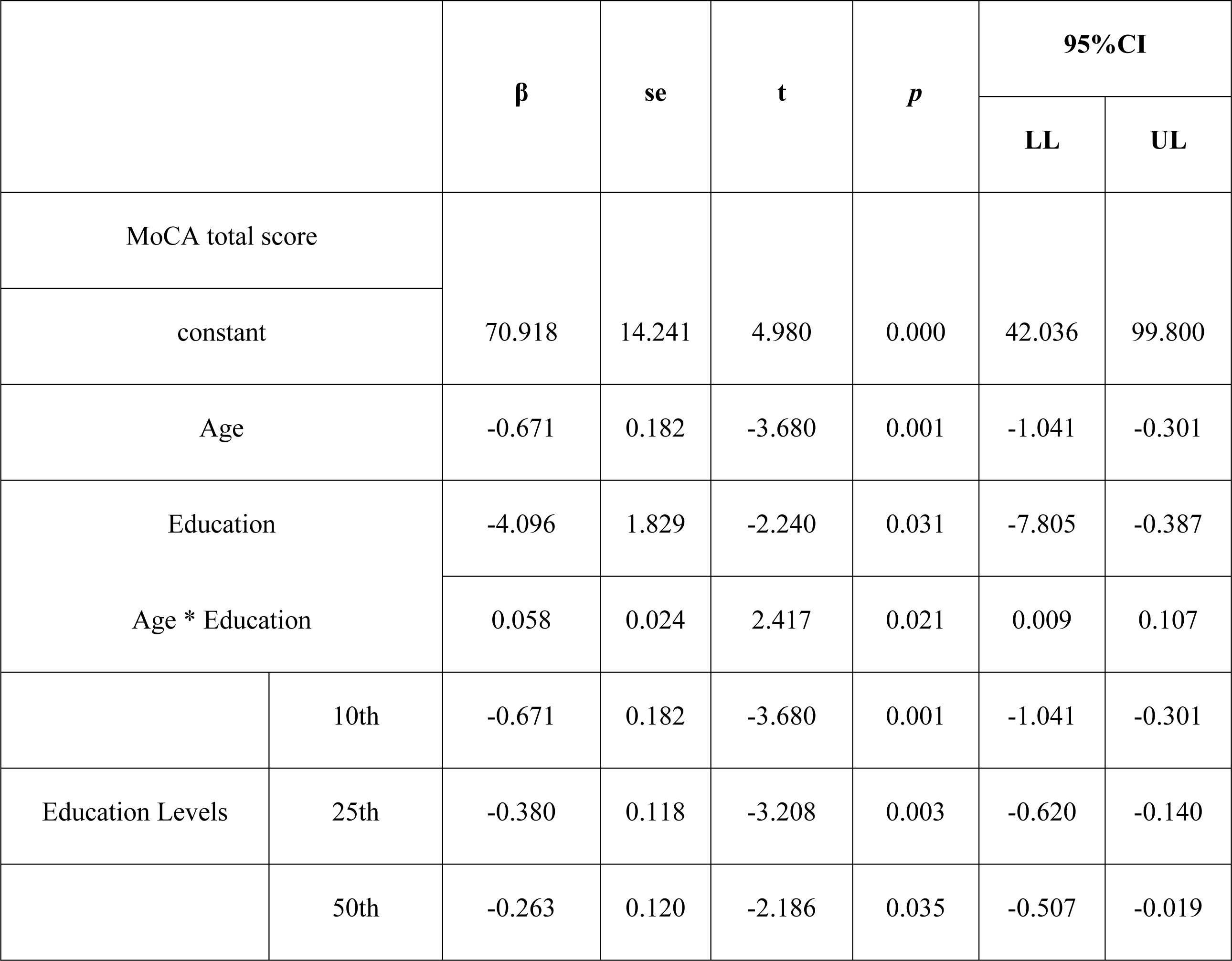
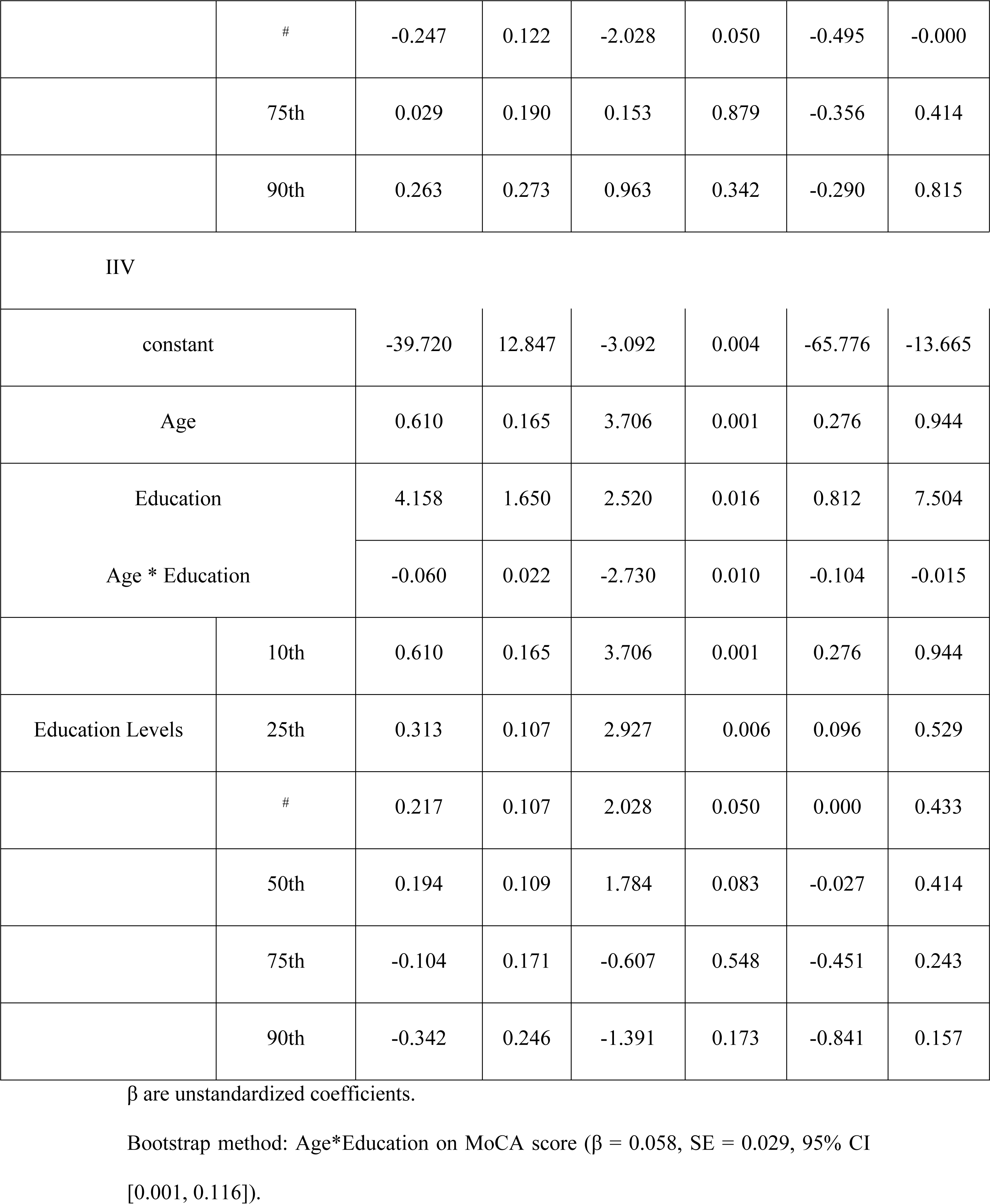
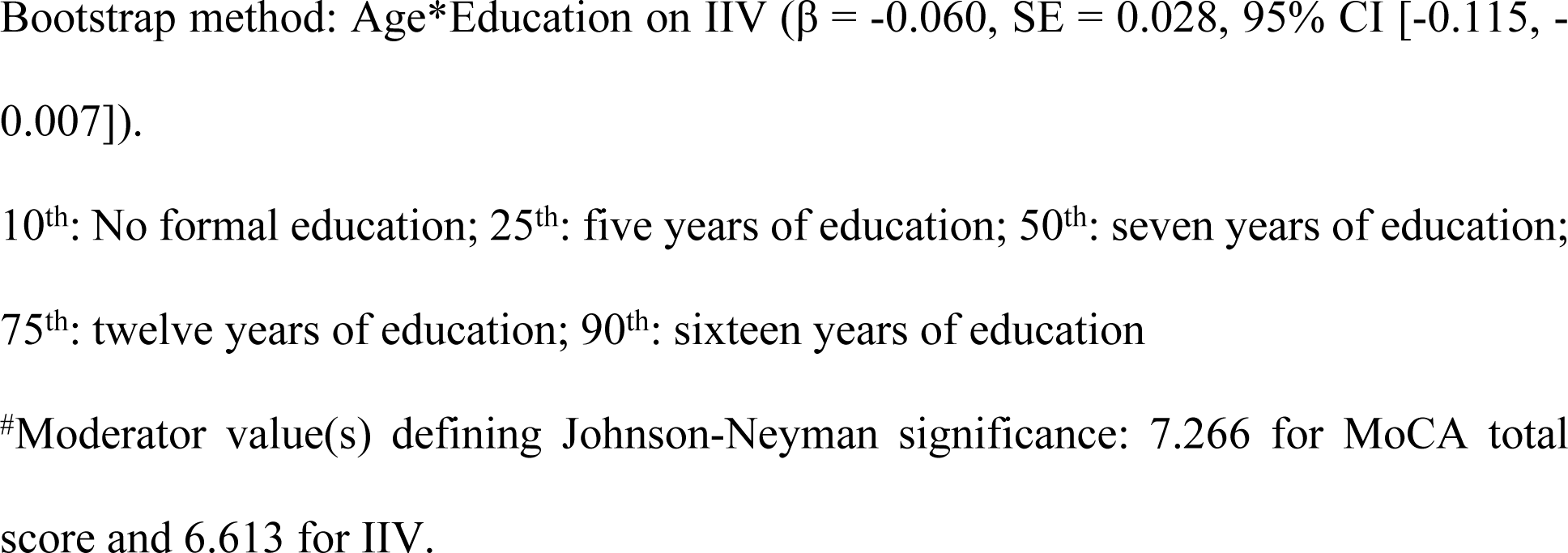
Line Regression Models for the Physically Frail Group.

In the PF group, for older adults who had no formal education, a negative association between age and the MoCA total score was observed (β = −0.671, SE = 0. 182, *p* = 0.001, 95% CI [−1.041, −0.301]; Fig 3A). For those with five years of education, we observed a significantly weaker negative association between age and MoCA total score (β = −0.380, SE = 0.118, *p* = 0.003, 95% CI [−0.620, −0.140]; Fig 3A). For those with 12 and 16 years of education, we observed a notable but non-statistically significant positive relationship between age and MoCA total score (β = 0.029, SE = 0.190, *p* = 0.879, 95% CI [−0.356, 0.414] and β = 0.263, SE = 0.273, *p* = 0.342, 95% CI [−0.290, 0.815] respectively; Fig 3A). Using the Johnson–Neyman technique, we found that the negative association between age and MoCA total score weakened as years of education increased. This association was not statistically significant when years of education exceeded seven years after adjusting for multiple comparions with Bonferroni correction (β = −0.247, SE = 0.122, *p* = 0.050, 95% CI [−0.495, −0.000]; Fig 3B). Similarly, after applying bootstrap estimation, we found a significant two-way interaction effect between education and age on MoCA total score (β = 0.058, SE = 0.029, 95% CI [0.001, 0.116]).

No significant moderation effect of education on the associations between age and MoCA total score was observed in the non-PF group (R-sq = 0.019, F = 1.507, *p* = 0.223).

### Moderation Effects of Education on the Association between Age and IIV

Years of education had a significant moderation effect on the association between age and IIV in the PF group. The overall model was statistically significant (R-sq = 0.512, F = 12.573, *p* < 0.001) (Table 2). Specifically, age exerted a significant positive main effect on IIV (i.e., older age was correlated with greater IIV) (β = 0.610, SE = 0.165, *p* = 0.001, 95% CI [0.276, 0.944]). We found a significant two-way interaction between education and age in the PF group (β = -0.060, SE = 0.022, *p* = 0.010, 95% CI [−0.104, −0.015]; Fig 4A) (Table 2), accounting for 10.110 % of the variance in IIV (F = 7.454, *p* = 0.010).

**Figure 4.**
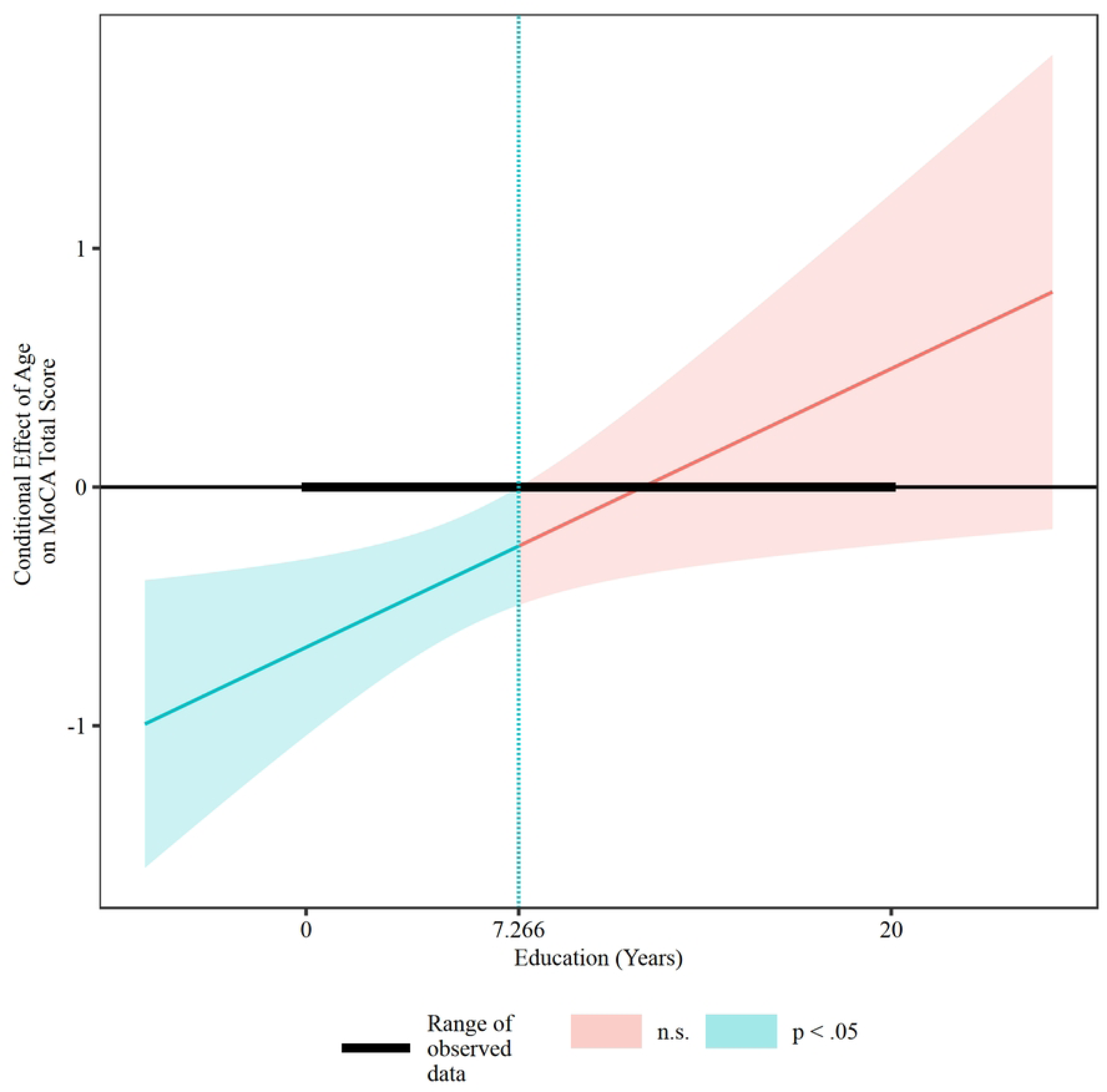
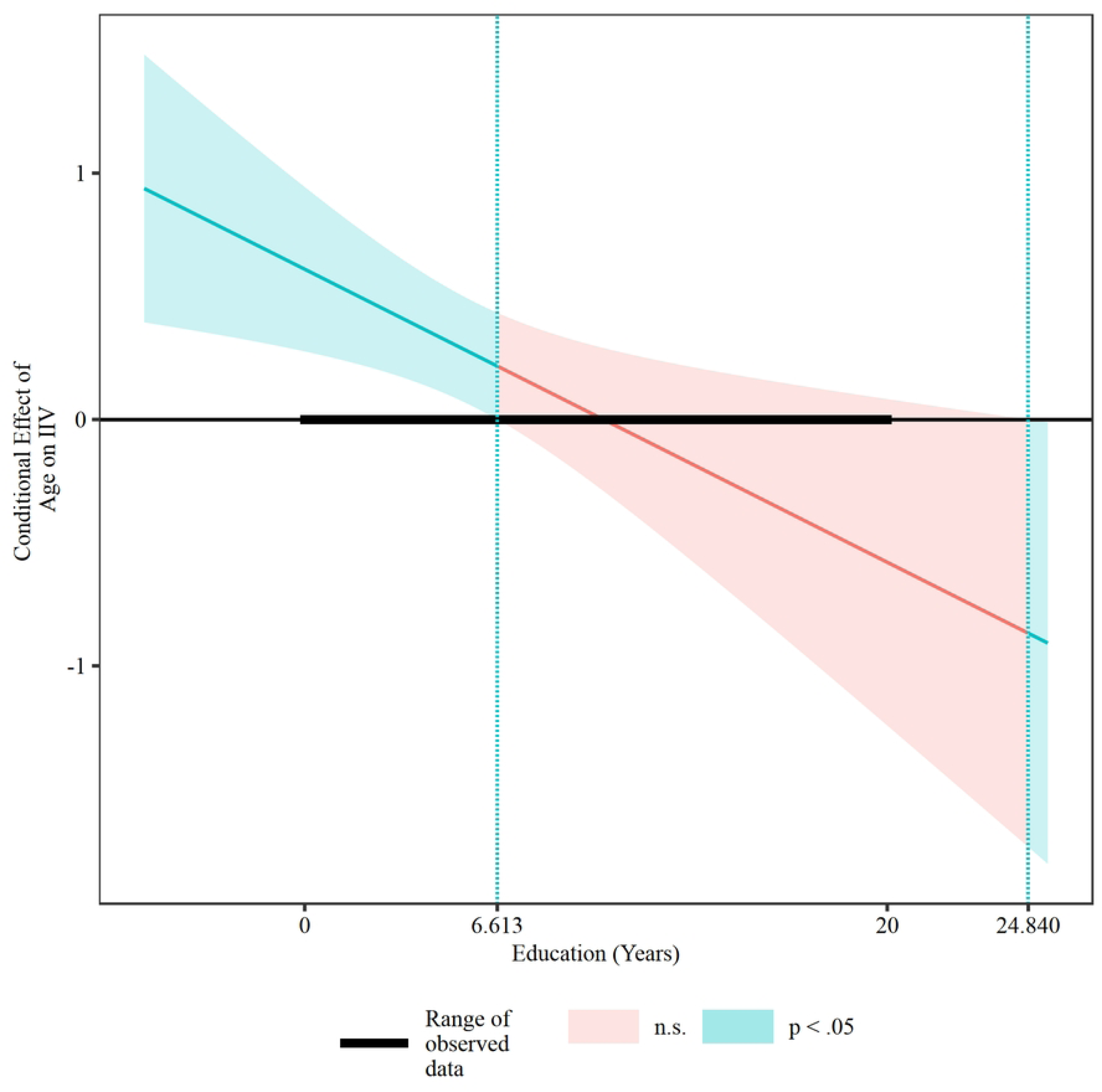
Moderation Effects of Education on the Association between age-IIV in the Physically Frail Group. (A) the association between age and IIV. (B) Jonnson-Neyman plot for visualizing the moderating effect of education between age and IIV.

In the PF group, for older adults who had no formal education, a positive association between age and IIV was observed (β = 0.610, SE = 0.165, *p* = 0.001, 95% CI [0.276, 0.944]; Fig 4A). For those with five years of education, we observed a significantly weaker positive association between age and IIV (β = 0.313, SE = 0.107, *p* = 0.006, 95% CI [0.096, 0.529]; Fig 4A); for those with 12 and 16 years of education, there was also a non-statistically significant negative association between age and IIV (i.e., older age was correlated with reduced IIV) (β = −0.104, SE = 0.171, *p* = 0.548, 95% CI [−0.451, 0.243] and β = −0.342, SE = 0.246, *p* = 0.173, 95% CI [−0.841, 0.157] respectively; Fig 4A). Using the Johnson–Neyman technique, we observed that the positive association between age and IIV weakened as years of education increased and the association was not statistically significant when years of education exceeded seven years after adjusting for multiple comparions with Bonferroni correction ((β = 0.217, SE = 0.107, *p* = 0.050, 95% CI [0.000, 0.430]; Fig 4B). Similarly, after applying bootstrap estimation, we found a significant two-way interaction effect between education and age on IIV (β = −0.060, SE = 0.028, 95% CI [−0.115, −0.007]).

No significant moderation effect of education on the associations between age and IIV was observed in the non-PF group (R-sq = 0.006, F = 0.439, *p* = 0.510).

## Discussion

In this cross-sectional study, we compared differences in IIV between older adults with and without physical frailty, and subsequently examined whether education level moderated the effects of aging on MoCA performance as assessed via MoCA total score as well as IIV. We found that compared with older adults without physical frailty, those with physical frailty demonstrated significantly lower MoCA total score and greater IIV. Additionally, our findings revealed that the cognitive benefits of education may vary depending on the health status of the individuals (i.e., with or without physical frailty); and whether cognitive function was evaluated as a static representation of their general cognition (i.e., MoCA total score), or as an index of sustained cognitive processing (i.e., IIV).

### Comparing Total Score between the PF and the non-PF Groups

Our findings revealed that compared with non-physically frail older adults, physically frail older adults had lower global cognitive function. This finding was in agreement with earlier studies (42, 43). For example, a cross-sectional that examined 4649 individuals aged 50 years and older indicated that physically frail individuals had worse MMSE and MoCA total scores compared with non-physically frail individuals (43).

### Comparing IIV between the PF and the non-PF Groups

We found that individuals with physical frailty showed greater IIV than those without physical frailty. These findings were in line with evidence documented in both healthy and cognitively impaired populations. Specifically, a study demonstrated that IIV, as indexed by dispersion across four cognitive domains (i.e., perceptual speed, semantic memory, episodic memory, and fluid reasoning) assessed by nine separate cognitive tests, can effectively discriminate between older adults with and without signs of cognitive decline (44). Halliday et al. (45) have also found that greater IIV as indexed by dispersion was associated with greater impairment in cognitive function across eight cognitive domains (i.e., attention, memory, executive function, processing speed, etc.) assessed by twelve separate cognitive tests among healthy older adults, older adults with MCI, and older adults with AD. Our results confirmed and extended these findings to the physically frail older population and showed that it may be possible to distinguish older adults with and without physical frailty via IIV calculated from a single cognitive test.

Notably, it is plausible that greater IIV displayed among PF older adults may be reflective of an impaired neural underpinnings of cognitive processing. A recent study suggested that greater IIV as indexed by dispersion was linked to neural noises - irregular neural activities that hinder cognitive processing - and reductions in the efficiency of neural information processing in the central nervous system (CNS) (46). Several neuroimaging studies indicated that the greater IIV as indexed by dispersion was notably associated with aberrant rest-state functional connectivity of the default mode network (DMN) (47, 48). For instance, greater intranetwork DMN connectivity was correlated with poorer dual-task performance; and greater connectivity between DMN and supplementary motor area was correlated with slower gait speed and greater postural sway in older adults with MCI (49). These findings further support the notion that IIV as indexed by dispersion may be a sensitive and promising indicator for cognitive decline and CNS dysfunction among older adults with physical frailty.

### Moderation Effects of Education on the Impact of Aging on IIV between the PF and the non-PF Groups

Early evidence suggested that the protective effects of education against cognitive decline may be dependent on physical frailty status. Specifically, study showed that compared with healthy older adults, physically frail older adults exhibited a stronger positive correlation between education and MMSE (50). Extending these findings, we found that education level moderated the association of age with sustained cognitive processing only in older adults with physical frailty. Education level is a well-established proxy of cognitive reserve (50, 51). Importantly, cognitive reserve describes an individual’s intrinsic capacity to withstand age-or disease-related pathologies and maintain cognitive function through efficient and effective adaptation of brain networks (52, 53). Stern (53, 54) proposed that older adults with higher cognitive reserve would be able to process cognitive tasks more efficiently, as the brain networks of those with high cognitive reserve were more adaptive, thereby these individuals were more capable in slowing aging-related cognitive decline when faced with the similar levels of age-or disease-related brain pathology. This was supported by neuroimaging studies. One study used graph theory compute human brain connectome and found that greater cognitive reserve was linked to greater global efficiency of brain networks (55). Additionally, by using years of education and scores of intelligence tests as proxies of cognitive reserve, Steffener et al. (56) found that cognitive reserve had a significantly indirect effect on memory performance through reducing the activation (i.e., greater neural efficiency) of the functional networks in older adults. These results suggest that older adults with higher education levels may have greater neural efficiency compared with those with lower education levels. Importantly, our findings aligned with a cross-sectional investigation that showed when compared with those with less education, older adults with 16 or more years of education displayed less IIV as indexed by dispersion (i.e., more stable, robust cognitive processing) (44).

Additionally, our results suggest that cognitive benefits from education may be more related to the ability to sustain robust cognitive processing (i.e., IIV), as opposed to a snapshot of the overall cognitive function (i.e., MoCA total score). This aligns well with previous study that administered 14 cognitive tests to older adults and only found notable differences in cognitive function indexed by IIV between the old and very older groups (57). It is plausible that education may mitigate impairments to sustained cognitive processing via maintaining neural efficiency of the neural networks that underpinned sustained cognitive processing. For instance, previous study found that education level was positively associated with local efficiency of brain networks by promoting more modular network configuration that is conducive to nodal communication and integration of information (55).

Notably, our findings suggest that the relationship between aging, cognitive function (i.e., MoCA total score and IIV), and education may be complex. Previous evidence also demonstrated the complex association between education and global cognitive function in older adults. Specifically, a longitudinal study that examined 260 older adults aged 60 years or older showed that eight years of education was linked to slower decline in MMSE score, but greater than nine years of education did not offer additional protection against cognitive decline (58). Similarly, in a cohort study, Mathuranath and colleagues (59) administered MMSE to 488 cognitively intact older adults and found that more than nine years of education did not offer further benefits in preventing cognitive decline. We extended these findings to physically frail older adults by reporting that years of education of more than 6.6 years did not further protect older physically frail individuals against decline in sustained cognitive processing. However, it is important to also note that while it was not statistically significant, the protective effects of education persisted beyond 6.6 years and were observed in those with more than 12 years, as well as 14 years with incremental increases in the protective effects. This suggests that there may be a ceiling effect in the obtainable cognitive benefits of education. Also, it is plausible that this ceiling effect may be population-dependent, as we observed significantly different moderation effects of education between those with and without physical frailty.

In the non-PF group, no significant moderation effect of education on the association between age and IIV was observed. It is probable that the non-PF group may exhibit greater brain reserve capacity. Brain reserve capacity is determined by brain structural integrity (53). The construct posited that individuals with greater brain reserve capacity have greater tolerance to pathologies, enabling the brain to have a higher threshold against insults (52). Hence, within the context of our findings, non-frail older adults may inherently possess greater brain reserve capacity such that the neuroprotective benefits of education were not required in these healthier individuals. Of note, our results align with previous study that reported no significant interaction effect between age and education on memory and general fluid intelligence among 603 healthy older adults over 70 (60). This evidence, in conjunction with our findings, suggests that attainable cognitive-protective effects of education may vary depending on the amount of available reserve capacity whereby the ceiling may be lower for those with a greater amount of reserve capacity. However, future studies will be needed to elucidate the relationship between physical frailty, brain reserve, and sustained cognitive processing.

The primary strength of our study is the novelty in using IIV computed from a single cognitive test to discriminate older adults with and without physical frailty. This may potentially reduce the time and effort required for clinical diagnosis. However, this study has several limitations. First, we enrolled physically frail older adults who were healthy enough to take part in research studies, therefore our findings cannot be generalized beyond this population. Second, some of the study participants included may be cognitively impaired, therefore we cannot rule out the potential confounding effects of mild cognitive impairment. Third, a single cognitive test was used to compute IIV, therefore our results cannot reflect sustained cognitive processing across multiple cognitive tests in this population. Additionally, longitudinal studies are needed to fully understand the impact of physical frailty on the trajectory of cognitive decline.

### Conclusion

This cross-section study provided evidence to support the use of IIV as a measure to identify physically frail older adults. Our findings also suggested that among older adults, the cognitive-protective benefits of education may be directly related to mitigating impaired capacity to sustain robust cognitive processing. However, the attainable protective effects of education may be dependent upon the overall health status of the older individuals.

## Institutional Review Board statement

This study received approval from the Institutional Review Board (IRB) of The Hong Kong Polytechnic University (HSEARS20230131001).

## Informed consent statement

Informed consent was obtained from all participants involved in the study.

## Data availability statement

All the data are available in this paper.

## Acknowledgments

The authors thank all participants for their valuable contributions.

## Authors Contribution

**Conceptualization:** Chun Liang HSU and Jingyi WU.

**Data curation:** Chun Liang HSU.

**Formal analysis:** Jingyi WU.

**Funding acquisition:** Chun Liang HSU.

**Investigation:** Chun Liang HSU, Jingyi WU, Jinyu Chen, and Juncen Wu.

**Methodology:** Jingyi WU.

**Project administration:** Chun Liang HSU.

**Resources:** Chun Liang HSU.

**Software:** Jingyi WU.

**Supervision:** Chun Liang HSU.

**Visualization:** Jingyi WU.

**Writing – original draft:** Jingyi WU.

**Writing – review & editing:** Chun Liang HSU

## References

1. Morley JE, Vellas B, Van Kan GA, Anker SD, Bauer JM, Bernabei R, et al. Frailty consensus: a call to action. Journal of the American Medical Directors Association. 2013;14(6):392–7.

2. O’Caoimh R, Sezgin D, O’Donovan MR, Molloy DW, Clegg A, Rockwood K, et al. Prevalence of frailty in 62 countries across the world: a systematic review and meta-analysis of population-level studies. Age and ageing. 2021;50(1):96–104.

3. Feng Z, Lugtenberg M, Franse C, Fang X, Hu S, Jin C, et al. Risk factors and protective factors associated with incident or increase of frailty among community-dwelling older adults: A systematic review of longitudinal studies. PloS one. 2017;12(6):e0178383.

4. Fried LP, Tangen CM, Walston J, Newman AB, Hirsch C, Gottdiener J, et al. Frailty in older adults: evidence for a phenotype. The Journals of Gerontology Series A: Biological Sciences and Medical Sciences. 2001;56(3):M146–M57.

5. Buchman AS, Boyle PA, Wilson RS, Tang Y, Bennett DA. Frailty is associated with incident Alzheimer’s disease and cognitive decline in the elderly. Psychosomatic medicine. 2007;69(5):483–9.

6. Ávila-Funes JA, Amieva H, Barberger-Gateau P, Le Goff M, Raoux N, Ritchie K, et al. Cognitive impairment improves the predictive validity of the phenotype of frailty for adverse health outcomes: the three-city study. Journal of the American Geriatrics Society. 2009;57(3):453–61.

7. Boyle PA, Buchman AS, Wilson RS, Leurgans SE, Bennett DA. Physical frailty is associated with incident mild cognitive impairment in community - based older persons. Journal of the American Geriatrics Society. 2010;58(2):248–55.

8. Mitnitski A, Fallah N, Rockwood M, Rockwood K. Transitions in cognitive status in relation to frailty in older adults: a comparison of three frailty measures. The journal of nutrition, health & aging. 2011;15:863–7.

9. Boyle PA, Buchman AS, Wilson RS, Leurgans SE, Bennett DA. Physical Frailty Is Associated with Incident Mild Cognitive Impairment in Community-Based Older Persons. Journal of the American Geriatrics Society (JAGS). 2010;58(2):248–55.

10. Nasreddine ZS, Phillips NA, Bédirian V, Charbonneau S, Whitehead V, Collin I, et al. The Montreal Cognitive Assessment, MoCA: a brief screening tool for mild cognitive impairment. Journal of the American Geriatrics Society. 2005;53(4):695-9.

11. Judge D, Roberts J, Khandker RK, Ambegaonkar B, Black CM. Physician practice patterns associated with diagnostic evaluation of patients with suspected mild cognitive impairment and Alzheimer’s disease. International Journal of Alzheimer’s Disease. 2019;2019.

12. Boker SM, Molenaar P, Nesselroade JR. Issues in intraindividual variability: individual differences in equilibria and dynamics over multiple time scales. Psychology and aging. 2009;24(4):858.

13. Hultsch DF, MacDonald SW. Intraindividual variability in performance as a theoretical window onto cognitive aging. New frontiers in cognitive aging. 2004:65–88.

14. Hultsch DF, Strauss E, Hunter MA, MacDonald SW. Intraindividual variability, cognition, and aging. The handbook of aging and cognition: Psychology Press; 2011. p. 497-562.

15. Christensen H, Mackinnon A, Korten A, Jorm A, Henderson A, Jacomb P. Dispersion in cognitive ability as a function of age: A longitudinal study of an elderly community sample. Aging, Neuropsychology, and Cognition. 1999;6(3):214–28.

16. MacDonald SW, Stawski RS. Intraindividual variability—an indicator of vulnerability or resilience in adult development and aging? Handbook of intraindividual variability across the life span: Routledge; 2014. p. 231–57.

17. Aita SL. Neuropsychological intra-individual variability: Review and meta-analysis in clinical adult samples: University of South Alabama; 2020.

18. Christensen H, Dear KB, Anstey KJ, Parslow RA, Sachdev P, Jorm AF. Within-occasion intraindividual variability and preclinical diagnostic status: is intraindividual variability an indicator of mild cognitive impairment? Neuropsychology. 2005;19(3):309.

19. Burton CL, Strauss E, Hultsch DF, Moll A, Hunter MA. Intraindividual variability as a marker of neurological dysfunction: a comparison of Alzheimer’s disease and Parkinson’s disease. Journal of Clinical and Experimental Neuropsychology. 2006;28(1):67–83.

20. Holtzer R, Verghese J, Wang C, Hall CB, Lipton RB. Within-person across-neuropsychological test variability and incident dementia. Jama. 2008;300(7):823–30.

21. Vaughan L, Leng I, Dagenbach D, Resnick SM, Rapp SR, Jennings JM, et al. Intraindividual variability in domain-specific cognition and risk of mild cognitive impairment and dementia. Current gerontology and geriatrics research. 2013;2013.

22. Brayne C, Ince PG, Keage HA, McKeith IG, Matthews FE, Polvikoski T, et al. Education, the brain and dementia: neuroprotection or compensation? EClipSE Collaborative Members. Brain. 2010;133(8):2210–6.

23. Brigola AG, Alexandre TdS, Inouye K, Yassuda MS, Pavarini SCI, Mioshi E. Limited formal education is strongly associated with lower cognitive status, functional disability and frailty status in older adults. Dementia & neuropsychologia. 2019;13:216–24.

24. Sharp ES, Gatz M. The relationship between education and dementia an updated systematic review. Alzheimer disease and associated disorders. 2011;25(4):289.

25. Alley D, Suthers K, Crimmins E. Education and cognitive decline in older Americans: Results from the AHEAD sample. Research on aging. 2007;29(1):73–94.

26. Cerreta F, Group EMAGE. New harmonized considerations on the evaluation instruments for baseline characterization of frailty in the European Union. British Journal of Clinical Pharmacology. 2020;86(10):2017–9.

27. Cerreta F, Ankri J, Bowen D, Cherubini A, Cruz Jentoft AJ, Guðmundsson A, et al. Baseline Frailty Evaluation in Drug Development. J Frailty Aging. 2016;5(3):139–40.

28. European Medicines Agency. Reflection paper on physical frailty: instruments for baseline characterisation of older populations in clinical trials. https://www.ema.europa.eu/documents/scientific-guideline/reflection-paper-physical-frailtyinstruments-baseline-characterisation-older-populations-clinical_en.pdf 2018 Accessed December 29, 2021.

29. Guralnik JM, Simonsick EM, Ferrucci L, Glynn RJ, Berkman LF, Blazer DG, et al. A short physical performance battery assessing lower extremity function: association with self-reported disability and prediction of mortality and nursing home admission. Journal of gerontology. 1994;49(2):M85–M94.

30. Guralnik JM, Ferrucci L, Pieper CF, Leveille SG, Markides KS, Ostir GV, et al. Lower extremity function and subsequent disability: consistency across studies, predictive models, and value of gait speed alone compared with the short physical performance battery. The Journals of Gerontology Series A: Biological Sciences and Medical Sciences. 2000;55(4):M221–M31.

31. da Câmara SMA, Alvarado BE, Guralnik JM, Guerra RO, Maciel ÁCC. Using the Short Physical Performance Battery to screen for frailty in young - old adults with distinct socioeconomic conditions. Geriatrics & gerontology international. 2013;13(2):421–8.

32. Bandinelli S, Lauretani F, Boscherini V, Gandi F, Pozzi M, Corsi AM, et al. A randomized, controlled trial of disability prevention in frail older patients screened in primary care: the FRASI study. Design and baseline evaluation. Aging clinical and experimental research. 2006;18:359–66.

33. Yeung P, Wong L, Chan C, Leung JL, Yung C. A validation study of the Hong Kong version of Montreal Cognitive Assessment (HK-MoCA) in Chinese older adults in Hong Kong. Hong Kong Medical Journal. 2014;20(6):504.

34. Yeung PY, Wong LL, Chan CC, Yung CY, Leung LJ, Tam YY, et al. Montreal cognitive assessment—single cutoff achieves screening purpose. Neuropsychiatric Disease and Treatment. 2020:2681–7.

35. Dawson B, Trapp RG. Basic & clinical biostatistics. Basic & clinical biostatistics 2004. p. 438-

36. Mascarenhas Fonseca L, Sage Chaytor N, Olufadi Y, Buchwald D, Galvin JE, Schmitter-Edgecombe M, et al. Intraindividual Cognitive Variability and Magnetic Resonance Imaging in Aging American Indians: Data from the Strong Heart Study. Journal of Alzheimer’s Disease. 2023(Preprint):1–13.

37. Yap B, Sim C. Comparison of various types of normality tests. Journal of Statistical Computation and Simulation. 2011;81(12):2141–55.

38. MacDonald SW, DeCarlo CA, Dixon RA. Linking biological and cognitive aging: toward improving characterizations of developmental time. Journals of Gerontology Series B: Psychological Sciences and Social Sciences. 2011;66(suppl_1):i59-i70.

39. Cohen J. Statistical power analysis for the behavioral sciences. 2nd ed. Hillsdale, N.J: L. Erlbaum Associates; 1988.

40. Hayes AF. Introduction to mediation, moderation, and conditional process analysis: A regression-based approach: Guilford publications; 2017.

41. Yzerbyt V, Muller D, Batailler C, Judd CM. New recommendations for testing indirect effects in mediational models: The need to report and test component paths. Journal of personality and social psychology. 2018;115(6):929.

42. Macuco CRM, Batistoni SST, Lopes A, Cachioni M, da Silva Falcão DV, Neri AL, et al. Mini-Mental State Examination performance in frail, pre-frail, and non-frail community dwelling older adults in Ermelino Matarazzo, São Paulo, Brazil. International psychogeriatrics. 2012;24(11):1725–31.

43. Robertson DA, Savva GM, Coen RF, Kenny RA. Cognitive function in the prefrailty and frailty syndrome. Journal of the American Geriatrics Society. 2014;62(11):2118–24.

44. Hilborn JV, Strauss E, Hultsch DF, Hunter MA. Intraindividual variability across cognitive domains: Investigation of dispersion levels and performance profiles in older adults. Journal of clinical and experimental neuropsychology. 2009;31(4):412–24.

45. Halliday DW, Stawski RS, Cerino ES, DeCarlo CA, Grewal K, MacDonald SW. Intraindividual variability across neuropsychological tests: Dispersion and disengaged lifestyle increase risk for Alzheimer’s disease. Journal of Intelligence. 2018;6(1):12.

46. Halliday DW, Gawryluk JR, Garcia-Barrera MA, MacDonald SW. White matter integrity is associated with intraindividual variability in neuropsychological test performance in healthy older adults. Frontiers in Human Neuroscience. 2019;13:352.

47. Mulet-Pons L, Solé-Padullés C, Cabello-Toscano M, Abellaneda-Pérez K, Perellón-Alfonso R, Cattaneo G, et al. Brain connectivity correlates of cognitive dispersion in a healthy middle-aged population: influence of subjective cognitive complaints. The Journals of Gerontology: Series B. 2023;78(11):1860–9.

48. Costa AS, Dogan I, Schulz JB, Reetz K. Going beyond the mean: Intraindividual variability of cognitive performance in prodromal and early neurodegenerative disorders. The Clinical Neuropsychologist. 2019;33(2):369–89.

49. Crockett RA, Hsu CL, Best JR, Liu-Ambrose T. Resting state default mode network connectivity, dual task performance, gait speed, and postural sway in older adults with mild cognitive impairment. Frontiers in aging neuroscience. 2017;9:423.

50. Ihle A, Gouveia ÉR, Gouveia BR, Freitas DL, Jurema J, Odim AP, et al. The relation of education, occupation, and cognitive activity to cognitive status in old age: the role of physical frailty. International psychogeriatrics. 2017;29(9):1469–74.

51. Farina M, Paloski LH, de Oliveira CR, de Lima Argimon II, Irigaray TQ. Cognitive reserve in elderly and its connection with cognitive performance: A systematic review. Ageing International. 2018;43:496–507.

52. Stern Y. Cognitive reserve in ageing and Alzheimer’s disease. The Lancet Neurology. 2012;11(11):1006–12.

53. Stern Y. What is cognitive reserve? Theory and research application of the reserve concept. Journal of the international neuropsychological society. 2002;8(3):448–60.

54. Stern Y, Habeck C, Moeller J, Scarmeas N, Anderson KE, Hilton HJ, et al. Brain networks associated with cognitive reserve in healthy young and old adults. Cerebral cortex. 2005;15(4):394–402.

55. Marques P, Moreira P, Magalhães R, Costa P, Santos N, Zihl J, et al. The functional connectome of cognitive reserve. Human brain mapping. 2016;37(9):3310–22.

56. Steffener J, Reuben A, Rakitin BC, Stern Y. Supporting performance in the face of age-related neural changes: testing mechanistic roles of cognitive reserve. Brain imaging and behavior. 2011;5:212–21.

57. Lindenberger U, Baltes PB. Intellectual functioning in old and very old age: cross-sectional results from the Berlin Aging Study. Psychology and aging. 1997;12(3):410.

58. Lyketsos CG, Chen L-S, Anthony JC. Cognitive decline in adulthood: an 11.5-year follow-up of the Baltimore Epidemiologic Catchment Area study. American Journal of Psychiatry. 1999;156(1):58–65.

59. Mathuranath P, Cherian JP, Mathew R, George A, Alexander A, Sarma SP. Mini mental state examination and the Addenbrooke’s cognitive examination: Effect of education and norms for a multicultural population. Neurology India. 2007;55(2):106–10.

60. Der G, Allerhand M, Starr JM, Hofer SM, Deary IJ. Age-related changes in memory and fluid reasoning in a sample of healthy old people. Aging, Neuropsychology, and Cognition. 2009;17(1):55–70.

